# Weighing the Hazards: Body Mass Index and Recurrence in Early-Onset Colorectal Cancer

**DOI:** 10.64898/2026.09.19.26363465

**Authors:** Erik Delryd, Andy Tran, Elinor Bexe Lindskog, David Ljungman

## Abstract

**Purpose:** Early-onset colorectal cancer (EOCRC, < 50 years) is rising sharply. The *obesity paradox*, where overweight correlates with better outcomes despite obesity being a risk factor, has been reported in colorectal cancer (CRC), mainly for overall survival, but its relevance to recurrence and to EOCRC specifically remains unclear. We investigated the association between body mass index (BMI) at diagnosis and risk of recurrence in EOCRC compared to average-onset colorectal cancer (AOCRC, ≥ 50 years).

**Methods:** This retrospective cohort study at Sahlgrenska University Hospital included 1821 patients who underwent radical surgery with curative intent for colorectal adenocarcinoma between 2011 and 2019, comprising 158 patients with EOCRC and 1663 with AOCRC. Cox models of recurrence were adjusted for tumour stage and location and stratified by differentiation. BMI was modelled with restricted cubic splines, and overall association and nonlinearity were assessed with likelihood-ratio tests (LRTs).

**Results:** In EOCRC, BMI was associated with recurrence (LRT p = 0.048), with a lower hazard among patients in the overweight range, although the test for nonlinearity was not significant (p = 0.08). In AOCRC, no significant association was observed (LRT p = 0.88). The interaction between BMI and cohort was not statistically significant (p = 0.11).

**Conclusion:** BMI at diagnosis was associated with recurrence in EOCRC but not in AOCRC. The observed pattern may be consistent with an obesity paradox in EOCRC. Larger studies incorporating prediagnostic weight and body composition are needed to confirm these findings.

## Introduction

Globally, a substantial increase in incidence of early-onset colorectal cancer (EOCRC; diagnosed before 50 years of age) has been recorded in recent decades [1]. The EOCRC incidence has increased the most among patients aged 20-29 with a 7.9% increase per year from 2004 to 2016 [2] and EOCRC is projected to become the leading cause of cancer-related death in patients aged 20-49 years by 2040 [3]. The reason for this increase is still to be established but studies have consistently shown that individuals with metabolic syndrome, characterised by conditions such as central obesity, hypertension, and dyslipidaemia, are at a significantly higher risk of developing colorectal cancer (CRC) [4]. CRC has also been linked to many environmental exposures, such as diets rich in processed meats [5], smoking [6], alcohol consumption [7, 8] and physical inactivity [9]. These associations have been noted for EOCRC as well [10-17].

Body mass index (BMI) has been extensively studied in relation to CRC risk and prognosis. Obesity is an established risk factor for the development of CRC [10, 18-21] and has also been linked to elevated risk of all-cause mortality, CRC-specific mortality, and recurrence in CRC patients of all ages [22-26]. Overweight, on the other hand, has been associated with a reduced risk of recurrence [21] and improved overall survival [25, 27-29]. This observation has been referred to in the literature as the *obesity paradox* [30]. Much of this evidence concerns overall survival, where part of the apparent benefit of excess weight may reflect lower non-cancer mortality, prediagnostic weight loss and selection [30, 31]. In a recent cohort of 5,521 patients, the survival advantage of overweight disappeared after adjustment for weight loss before diagnosis [32], and a nationwide Danish cohort of patients with stage I-III CRC found no association between BMI and recurrence [33].

The obesity paradox has mainly been described in patients with CRC irrespective of age [21, 25, 27-30], and few studies have examined the association between BMI and outcome in EOCRC specifically [34]. The aim of this study was to examine the association between BMI at diagnosis and recurrence after radical surgery in EOCRC, and to compare it with average-onset colorectal cancer (AOCRC). Recurrence was here selected as the endpoint because it more directly reflects the course of the cancer after curative treatment and is less directly affected by deaths from other causes than overall survival, particularly in older patients.

## Methods

### Data Source

This was a single-institution retrospective cohort study of patients treated for colorectal cancer at Sahlgrenska University Hospital between 2011 and 2019. Electronic health records were reviewed for demographic, clinical, and outcome characteristics.

### Study Population

Patients who underwent curative-intent surgery for histologically confirmed colorectal adenocarcinoma between 2011 and 2019 were included. The study period was restricted to 2011-2019 because height and weight data and adequate follow-up were available during this time. Reasons for exclusion were resections classified as non-radical or of uncertain radicality, missing height or weight, missing model covariates, or missing recurrence status. Patients with BMI above 40 (EOCRC n = 1, AOCRC n = 14) were also excluded because too few patients had values in this range for the spline to be estimated reliably.

### Variables

Patient characteristics included age, sex, and BMI. BMI was calculated from height and weight measured during the initial diagnostic workup, before neoadjuvant treatment, and categorised based on the World Health Organization’s guidelines [35] as underweight (BMI < 18.5), normal-weight (18.5 ≤ BMI < 25), overweight (25 ≤ BMI < 30), and obese (BMI ≥ 30). Clinicopathological characteristics included cancer diagnosis (colon, rectal, or both), tumour location (right-sided, left-sided, or rectum), tumour stage, T and N classification, and differentiation (poor, moderate, well, or mucinous). Tumour stage was classified according to the AJCC staging system, using pathological staging following surgery (pTNM) or post-treatment pathological staging (ypTNM) for patients who received neoadjuvant treatment. Distant metastases were classified using available clinical and radiological findings. Histopathological features such as tumour perforation, perineural growth, vascular growth, and EMVI were reported as binary variables. The number of lymph nodes examined and the number positive for metastases were also recorded. Treatment characteristics included neoadjuvant and adjuvant chemotherapy or radiotherapy, type of surgery (e.g., right-sided, left-sided, rectal resection or colectomy) along with surgical modality (elective, laparoscopic, radical, metastasis resection). Outcomes included new CRC, recurrence, and vital status at the time of last follow-up. Time to recurrence, death, and last follow-up were calculated from the date of radical surgery with curative aim, defined as the primary operation or, when radicality was achieved through additional surgery, the date of that operation. The primary endpoint was time to recurrence, defined as the interval from radical surgery to the first documented local or distant recurrence or death from colorectal cancer, whichever occurred first. Deaths from other causes were censored at the date of death. All deaths from colorectal cancer were preceded by a documented recurrence. Median follow-up was estimated with the reverse Kaplan-Meier method.

### Statistical Analysis

Continuous variables are presented as means with standard deviations (SD), and categorical variables as frequencies or proportions. Differences across BMI categories were tested with ANOVA for continuous variables and with Fisher’s exact test (EOCRC) or the chi-squared test (AOCRC) for categorical variables. The number of missing values for variables not included in the models is reported in eTables 1 and 2.

**Table 1.** Hazard ratios for recurrence by BMI category in EOCRC and AOCRC.

| BMI category | n / events | Unadjusted HR (95% CI) | p | Adjusted HR (95% CI)<sup>a</sup> | p |
|---|---|---|---|---|---|
| **EOCRC** |  |  |  |  |  |
| Underweight | 3 / 1 | 1.42 (0.19-10.57) | 0.73 | 0.73 (0.09-5.50) | 0.73 |
| Normal-weight | 79 / 26 | Reference |  | Reference |  |
| Overweight | 61 / 10 | 0.41 (0.20-0.86) | 0.02 | 0.53 (0.23-1.19) | 0.12 |
| Obese | 15 / 3 | 0.48 (0.16-1.59) | 0.23 | 0.45 (0.15-1.37) | 0.16 |
| **AOCRC** |  |  |  |  |  |
| Underweight | 32 / 6 | 1.31 (0.57-2.98) | 0.51 | 1.77 (0.77-4.03) | 0.18 |
| Normal-weight | 744 / 123 | Reference |  | Reference |  |
| Overweight | 627 / 109 | 1.07 (0.82-1.38) | 0.59 | 0.95 (0.74-1.24) | 0.75 |
| Obese | 260 / 47 | 1.09 (0.78-1.53) | 0.60 | 1.11 (0.79-1.56) | 0.52 |

<sup>a</sup>Adjusted for tumour location and stage and stratified by differentiation.
BMI categories: underweight < 18.5, normal-weight 18.5-24.9, overweight 25-29.9 and obese 30-40. Normal-weight is the reference category.
BMI, body mass index; HR, hazard ratio; CI, confidence interval; EOCRC, early-onset colorectal cancer; AOCRC, average-onset colorectal cancer

**Table 2.** Hazard ratios for recurrence by covariates in EOCRC and AOCRC.

| Variable | EOCRC |  | AOCRC |  |
|---|---|---|---|---|
|  | HR (95% CI) | p | HR (95% CI) | p |
| **Tumour location** |  |  |  |  |
| Right colon | Reference |  | Reference |  |
| Left colon | 0.42 (0.16-1.11) | 0.08 | 0.92 (0.68-1.26) | 0.61 |
| Rectum | 0.76 (0.37-1.56) | 0.46 | 1.19 (0.89-1.58) | 0.22 |
| **Stage** |  |  |  |  |
| I or II | Reference |  | Reference |  |
| III | 4.07 (1.49-11.1) | 0.006 | 4.56 (3.36-6.21) | < 0.001 |
| IV | 13.1 (4.41-38.9) | < 0.001 | 16.93 (11.78-24.37) | < 0.001 |

Hazard ratios from the adjusted model in Table 1. Models were stratified by differentiation, so no hazard ratio is estimated for it.
HR, hazard ratio; CI, confidence interval; EOCRC, early-onset colorectal cancer; AOCRC, average-onset colorectal cancer

Associations between BMI and time to recurrence were examined using multivariable Cox proportional hazards regression adjusted for tumour location (right-sided, left-sided, rectum), stage (I-II, III, IV), and stratified by differentiation. Complete pathological response after neoadjuvant treatment (ypT0N0) was grouped with stage I-II. Covariates were selected a priori as established prognostic factors. Treatment variables were not included. Neoadjuvant treatment is decided on the basis of clinical stage before treatment, which differs from the post-treatment stage used in the model. Neoadjuvant treatment was instead addressed in a sensitivity analysis excluding patients who received treatment expected to cause downstaging.

The proportional hazards assumption was assessed with Schoenfeld residuals. Because differentiation violated the assumption, models were stratified by differentiation, which allows a separate baseline hazard for each category without adding parameters.

BMI was modelled with restricted cubic splines with four knots at the 5th, 35th, 65th and 95th percentiles of BMI within each cohort, as recommended by Harrell [36]; knot locations are given in the figure legends. Spline models were adjusted for tumour location and stage and stratified by differentiation. The combined model used knots from the combined BMI distribution and was also stratified by cohort. A BMI of 21.75, the midpoint of the normal weight range, was set as the reference value. Curves were plotted between the 5th and 95th percentiles of BMI, to avoid unstable estimates at the extremes of the BMI distribution.

Likelihood-ratio tests (LRTs) were used to assess whether inclusion of BMI improved model fit within each cohort. Nonlinearity was assessed with an LRT comparing the spline model with a model in which BMI was entered as a linear term. Interaction between BMI and the cohorts was assessed in the combined cohort, with knots at the percentiles of the combined BMI distribution. The spline model was prespecified as the primary analysis, alongside a categorical BMI analysis.

The primary model had seven parameters (three for BMI, two for tumour location and two for stage), giving 5.7 events per parameter in EOCRC. This is below the ten events per variable often recommended [37], but within the range of five to nine events per variable in which simulation studies have found acceptable performance [38]. To assess whether the results depended on model size, BMI was also entered as a single linear term in additional robustness analyses, and reported as the HR per 5 kg/m^2^ increase in BMI, a scale commonly used in studies of BMI and cancer [39]. These models included BMI with tumour location and stage (five parameters), BMI with colon vs rectum and stage (four parameters), and the model with tumour location and stage was repeated in patients with stage I-III only.

All analyses were conducted using Stata version 18.0 (StataCorp LLC, College Station, TX, USA).

## Results

### Patient characteristics

The study included 158 patients with EOCRC and 1,663 patients with AOCRC, and the mean age at surgery was 43.0 ± 5.7 years in EOCRC and 71.1 ± 9.7 years in AOCRC. The proportion of men was similar in both cohorts (52.5% and 53.5%, respectively). In EOCRC, 3 patients (1.9%) were underweight, 79 (50.0%) normal-weight, 61 (38.6%) overweight and 15 (9.5%) obese. The corresponding numbers in AOCRC were 32 (1.9%), 744 (44.7%), 627 (37.7%) and 260 (15.6%).

Colon cancer accounted for 57.0% of EOCRC and 60.9% of AOCRC cases. Advanced-stage disease (stage III-IV) was more common in EOCRC than in AOCRC (60.1% vs 44.4%), as was the use of adjuvant chemotherapy (69.6% vs 36.7%). Most procedures were elective in both cohorts (96.2% and 95.8%, respectively). Median follow-up was 4.8 years in EOCRC and 3.5 years in AOCRC. Recurrence occurred in 40 (25.3%) and 285 (17.1%) patients, respectively.

Among all operated patients, BMI data were missing for 24.5% of EOCRC and 21.5% of AOCRC patients. Compared with patients with BMI data, those without more often underwent emergency surgery (50.9% vs 5.1% in EOCRC; 40.3% vs 4.9% in AOCRC), had stage IV disease (52.7% vs 18.9%; 37.0% vs 13.7%) and underwent non-radical surgery (45.6% vs 8.0%; 32.6% vs 9.4%). Among radically operated patients, recurrence rates were similar in patients without and with BMI data (25.8% vs 25.5% in EOCRC, p = 0.99; 19.5% vs 17.0% in AOCRC, p = 0.27).

### BMI and recurrence

In EOCRC, recurrence was observed in 26 of 79 normal-weight patients (32.9%), compared with 10 of 61 overweight patients (16.4%) and 3 of 15 obese patients (20.0%). One of the three underweight patients experienced recurrence. The estimated five-year probability of remaining recurrence-free was 60.5% for normal-weight patients, 83.0% for overweight patients, and 78.6% for obese patients. In AOCRC, corresponding estimates ranged from 74.4% to 76.6% across BMI categories. Kaplan-Meier curves are shown in eFigures 1 and 2. The site of recurrence after radical surgery by BMI category is shown in eTable 3.

**Fig 1.**
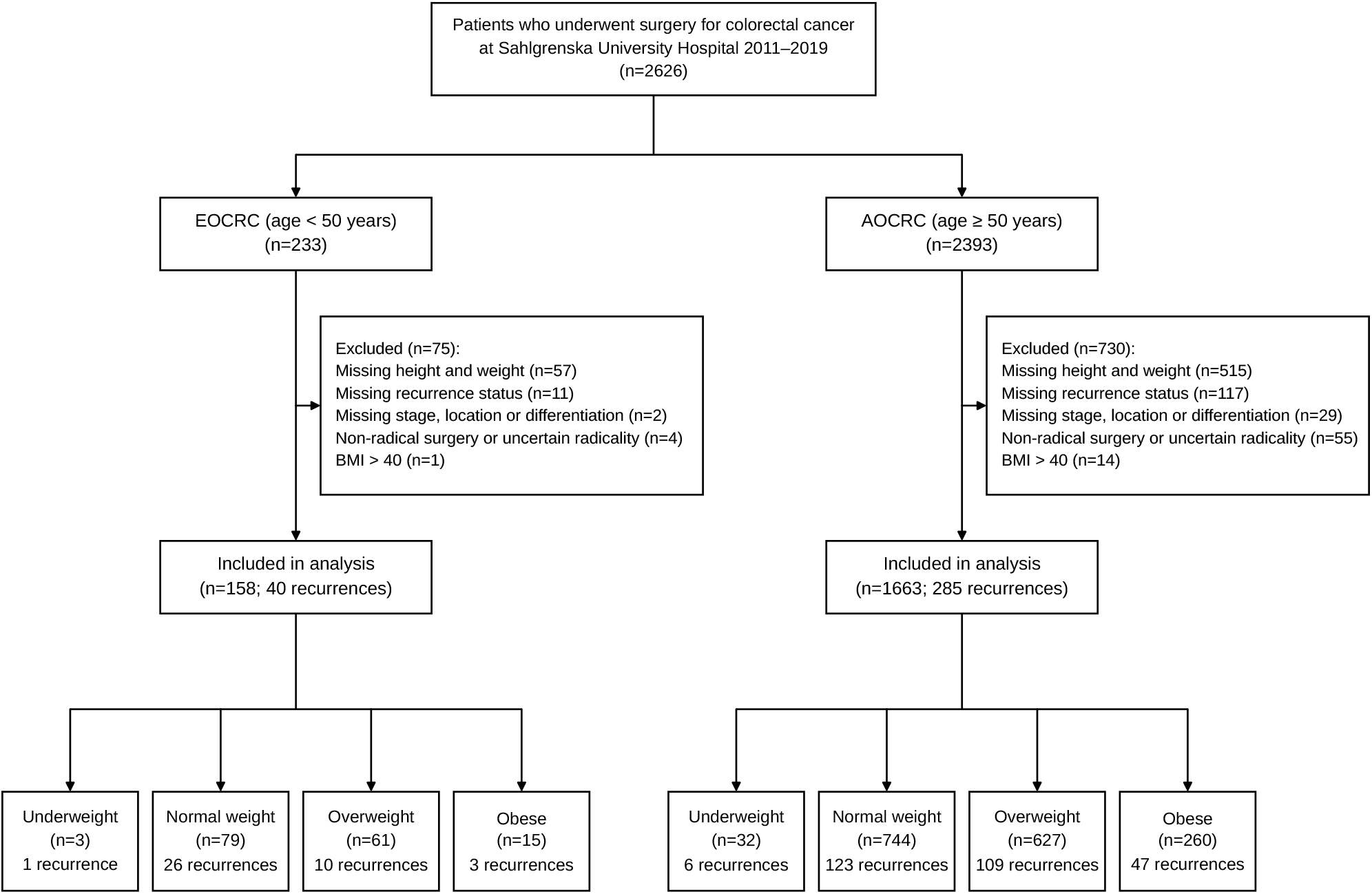
Flowchart of inclusion process

**Fig 2.**
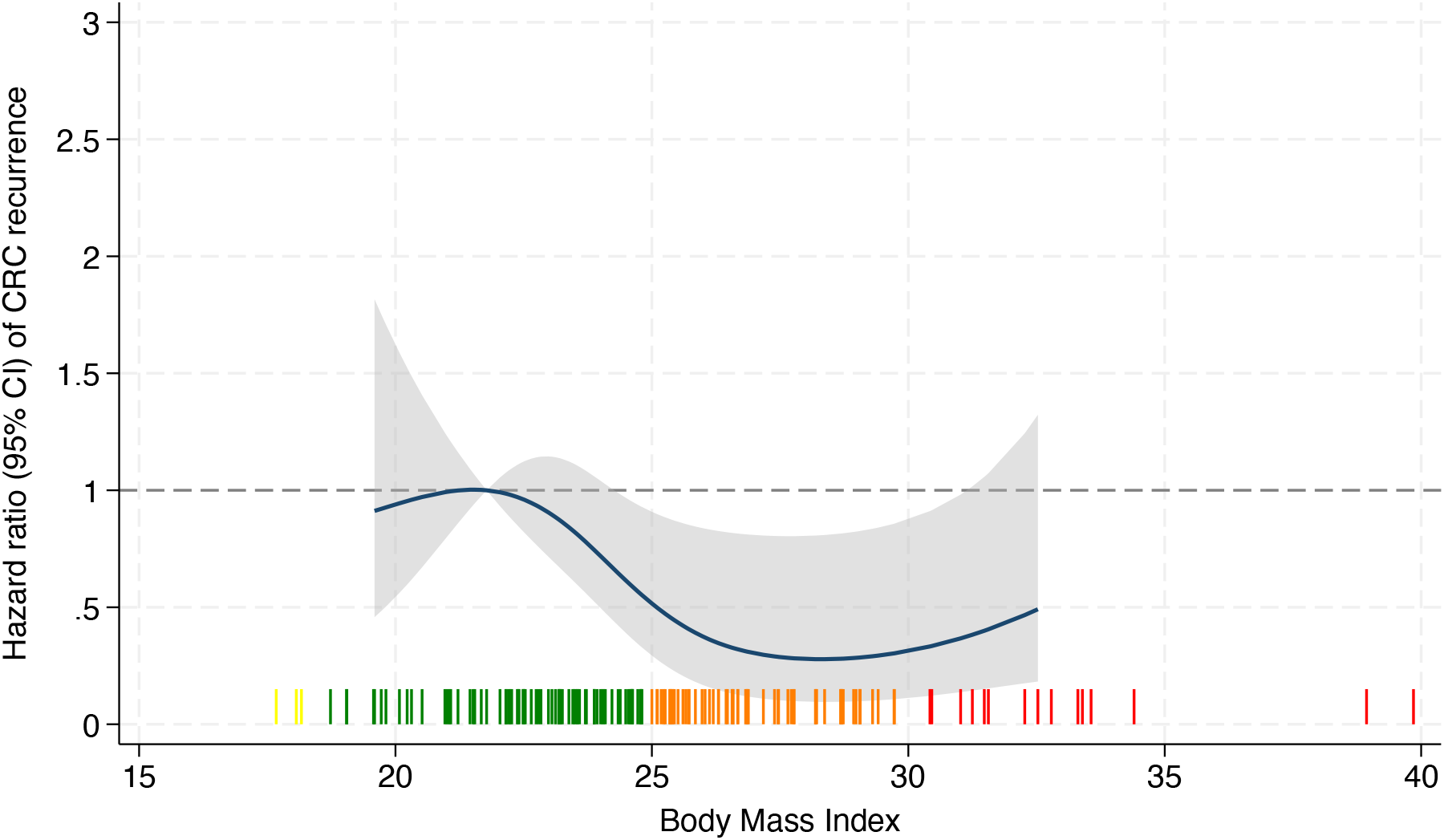
Hazard ratios for recurrence by BMI in EOCRC (n = 158; 40 recurrences). Knots at BMI 19.59, 23.57, 26.15 and 32.54. The shaded area represents the 95% confidence interval, and the rug plot shows individual patients by BMI category (yellow, underweight; green, normal weight; orange, overweight; red, obese).

**Table 3.** Robustness and sensitivity analyses for the association between BMI and recurrence.

| Model | EOCRC |  |  |  |
|---|---|---|---|---|
|  | Parameters | n / events | HR (95% CI) | p |
| **Model Size** |  |  |  |  |
| 1. Spline + location + stage (primary) | 7 | 158 / 40 | -<sup>a</sup> | 0.048<sup>b</sup> |
| 2. Categorical BMI + location + stage | 7 | 158 / 40 | -<sup>c</sup> | 0.28 |
| 3. Linear BMI + location + stage | 5 | 158 / 40 | 0.67 (0.43-1.07) | 0.09 |
| **Sensitivity analyses** |  |  |  |  |
| Model 3, excluding neoadjuvant treatment<sup>d</sup> | 5 | 125 / 27 | 0.77 (0.41-1.44) | 0.41 |
| Model 3, divided colon/rectum | 4 | 158 / 40 | 0.71 (0.45-1.13) | 0.15 |
| Model 3, Stage I-III only | 4 | 141 / 27 | 0.65 (0.37-1.14) | 0.11 |

All models are stratified by differentiation, which adds no parameters. Model 3 includes BMI as a linear term.
<sup>a</sup>No single hazard ratio: BMI is modelled with three spline terms.
<sup>b</sup>Likelihood-ratio test of the overall association. Test for nonlinearity: p = 0.08.
<sup>c</sup>Hazard ratios for each category are given in Table 1, p is the likelihood-ratio test for all categories.
<sup>d</sup>Patients who received long-course radiotherapy with or without chemotherapy, short-course radiotherapy with delayed surgery, or neoadjuvant chemotherapy were excluded.
HR, hazard ratio; CI, confidence interval; EOCRC, early-onset colorectal cancer

When BMI was modelled continuously using restricted cubic splines, BMI was associated with recurrence in the EOCRC cohort (LRT p = 0.048). Relative to the reference BMI of 21.75, the pointwise 95% confidence intervals for the HR were below 1 between BMI values of approximately 25 and 30, although the test for nonlinearity was not statistically significant (p = 0.08) (Fig. 2). In AOCRC, the spline curve showed no evidence of an overall association (LRT p = 0.88) or of nonlinearity (p = 0.91) (Fig. 3). In the combined cohort, there was similarly no evidence of an association (Fig. 4).

**Fig 3.**
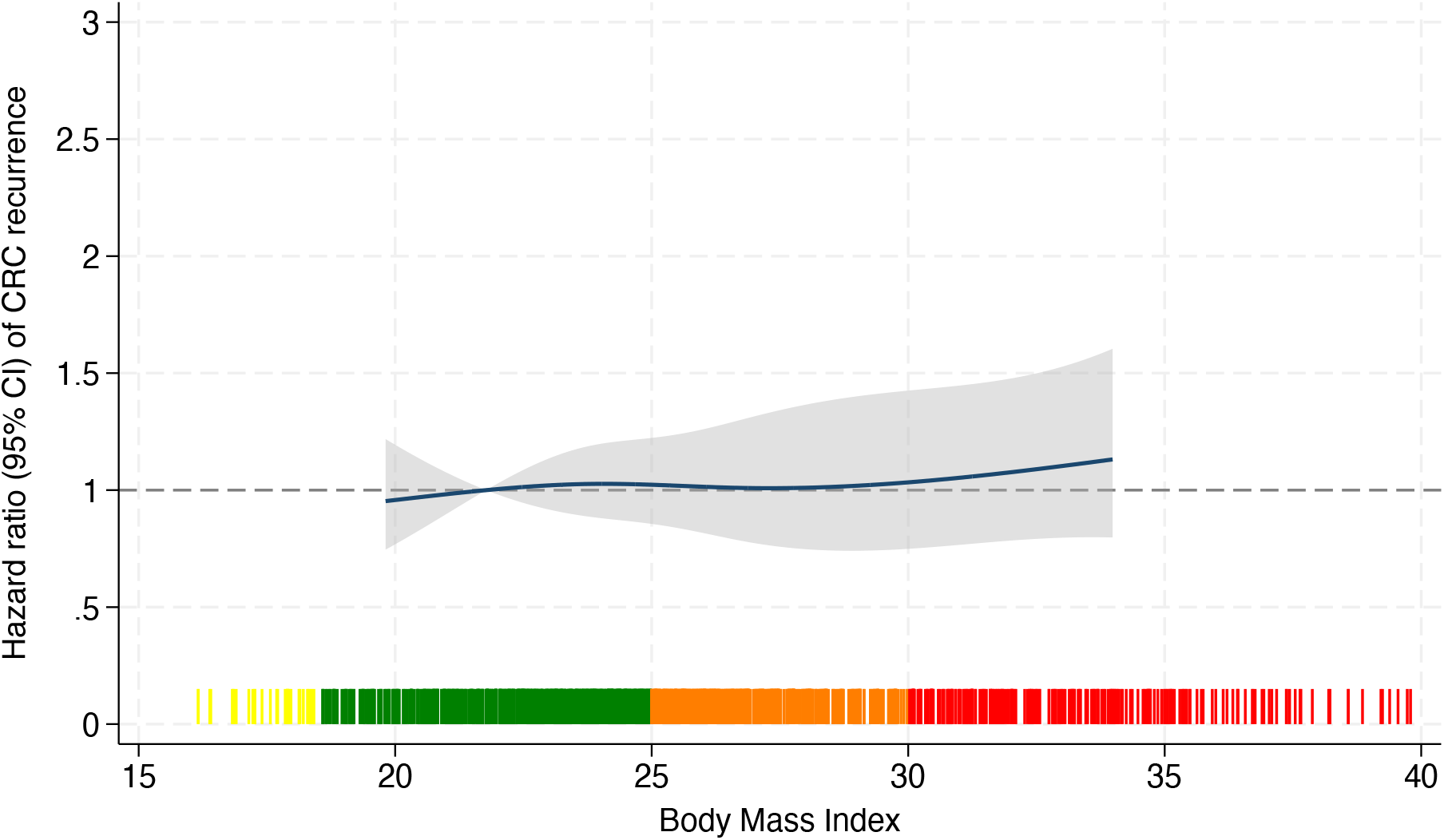
Hazard ratios for recurrence by BMI in AOCRC (n = 1,663; 285 recurrences). Knots at BMI 19.79, 23.95, 26.79 and 34.01. The shaded area represents the 95% confidence interval, and the rug plot shows individual patients by BMI category (yellow, underweight; green, normal weight; orange, overweight; red, obese).

**Fig 4.**
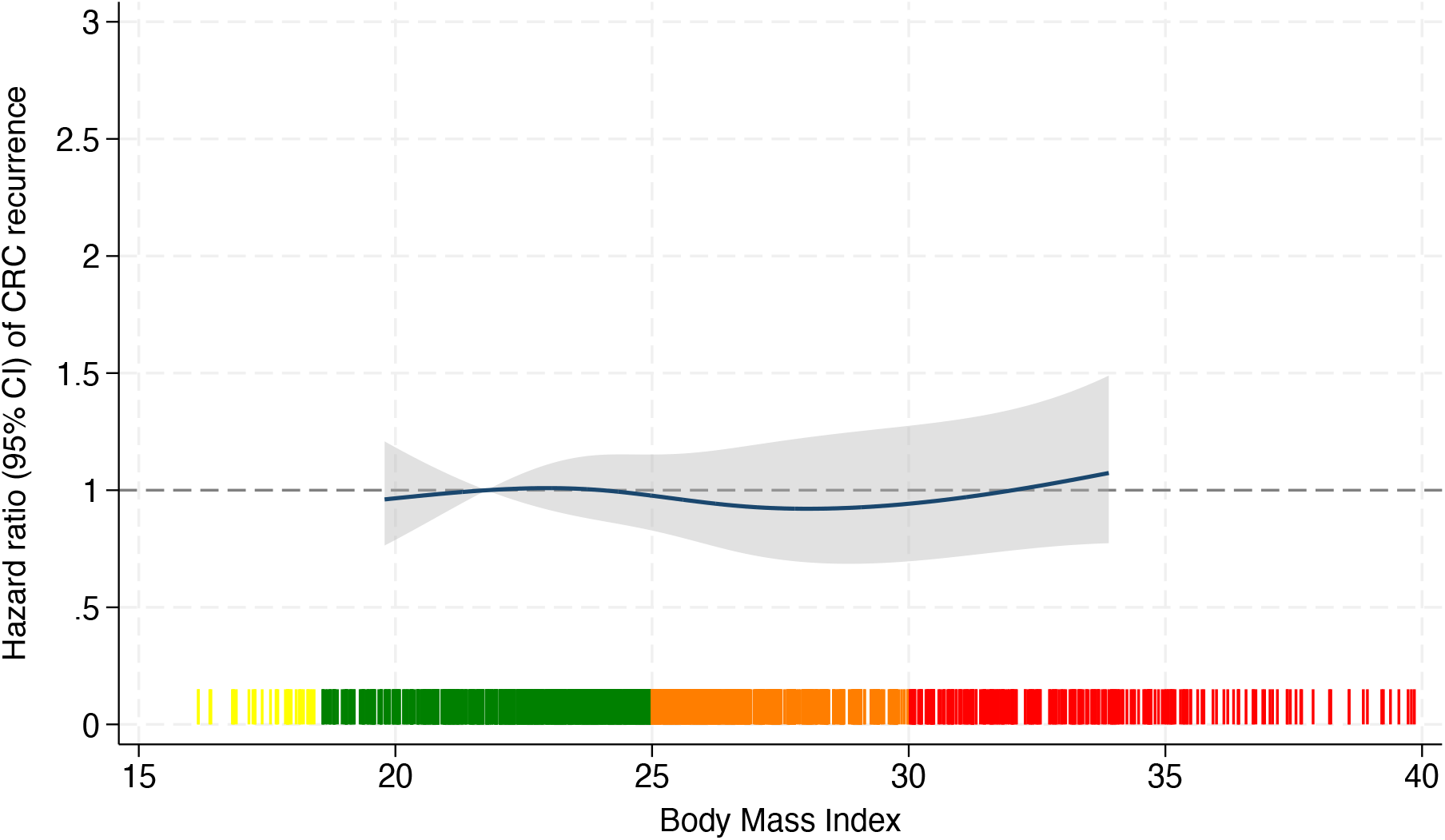
Hazard ratios for recurrence by BMI in the combined cohort (n = 1,821; 325 recurrences). Knots at BMI 19.72, 23.92, 26.73 and 33.94. The shaded area represents the 95% confidence interval, and the rug plot shows individual patients by BMI category (yellow, underweight; green, normal weight; orange, overweight; red, obese).

The categorical Cox analyses showed a similar pattern in EOCRC. Compared with normal-weight patients, overweight patients had a lower hazard of recurrence in the unadjusted analysis (HR 0.41, 95% CI 0.20-0.86, p = 0.02). After adjustment for tumour location and stage, the association was no longer statistically significant (HR 0.53, 95% CI 0.23-1.19, p = 0.12). No statistically significant associations were observed for the other BMI categories in EOCRC or for any BMI category in AOCRC (Table 1). Although the spline curves differed visually between cohorts, interaction tests did not provide evidence of a statistically significant difference, either with BMI modelled using splines (LRT p = 0.11) or as a linear term (p = 0.12).

### Robustness and sensitivity analyses

Assessment of the proportional hazards assumption using Schoenfeld residuals indicated violations for tumour differentiation in both cohorts, specifically for moderately or well-differentiated tumours in EOCRC (p = 0.005) and mucinous tumours in AOCRC (p = 0.006). The Cox models were therefore stratified by tumour differentiation.

In EOCRC, the hazard was lower at higher BMI in the linear model, but the association was not statistically significant (HR per 5 kg/m^2^ 0.67, 95% CI 0.43-1.07, p = 0.09). The results were similar in models with fewer parameters and after excluding patients with stage IV disease or neoadjuvant treatment expected to cause downstaging (Table 3).

## Discussion

In this study, BMI at diagnosis was associated with recurrence in EOCRC but not in AOCRC. The spline analysis suggested a lower recurrence risk in the overweight BMI range, although the test for nonlinearity was inconclusive. A similar association was observed in the categorical analysis, but the association was attenuated and no longer statistically significant after adjustment for tumour location and stage. However, the interaction between BMI and cohort was not statistically significant. The observed pattern may be consistent with an obesity paradox in EOCRC, potentially reflecting differences in body composition, weight loss patterns, tumour biology or systemic metabolism between younger and older patients. One possible explanation for the observed association in EOCRC is reverse causation, whereby patients with more advanced disease may have lost weight before diagnosis, resulting in a lower BMI at diagnosis. In our cohort, stage IV disease was present in 16.5% of normal-weight EOCRC patients compared with 1.6% of overweight patients, consistent with this explanation. In AOCRC, the proportion was similar across BMI groups (6-7%), possibly because older patients with metastatic disease and weight loss are less often offered radical surgery, which was required for inclusion. Since BMI was measured only once, at diagnosis, weight loss before diagnosis could not be accounted for. Previous studies have shown that the obesity paradox in CRC may disappear when weight loss before diagnosis is taken into account [30, 32, 40].

The observed pattern in EOCRC may be consistent with an obesity paradox, although our findings do not establish a protective effect of higher BMI. Previous studies have reported better outcomes among overweight patients with CRC [25, 27-30, 40], and lower risk of recurrence [21]. Our findings differ in part from those of a previous single-centre study of EOCRC of similar size, which found no significant difference in disease-free survival across BMI categories, although obesity was associated with worse overall and cancer-specific survival [34]. That study included a larger proportion of patients with obesity and used disease-free survival as the endpoint. In AOCRC, our findings agree with the Danish nationwide cohort study in which BMI was not associated with recurrence [33]. Although previous studies have examined BMI and CRC prognosis, relatively few have specifically investigated recurrence in EOCRC. Our study adds to this literature by examining BMI both categorically and continuously in EOCRC and AOCRC, while also assessing whether the association differs between cohorts.

Another possible explanation is differences in body composition. BMI does not distinguish between muscle mass and fat mass, and patients with similar BMI values may have very different body compositions. Younger patients with higher BMI may have higher muscle mass than older patients [41], while older patients are more likely to have sarcopenia and increased visceral fat, both associated with poorer outcomes [28, 40]. CT-based studies have shown considerable variation in muscle mass in patients within the same BMI category [42], and the prevalence of muscle mass loss in patients over 80 years has been reported to be as high as 50% [43]. However, body composition was not measured in this study, and these hypotheses cannot be tested with our data.

Overall, BMI was associated with recurrence in EOCRC, although the shape and strength of the association remain uncertain. The findings should be confirmed in larger cohorts with information on prediagnostic weight and body composition.

This study has several strengths. Both cohorts were drawn from the same institutional database, with similar data collection and definitions. Recurrence was used as the primary endpoint, and BMI was analysed using restricted cubic splines, linear Cox models, and BMI categories. Restricted cubic splines allowed us to examine possible nonlinear associations without assuming a linear relationship. Several robustness and sensitivity analyses were also performed.

The main limitation was the relatively small EOCRC cohort, with only 40 recurrences. The primary Cox model included seven parameters, corresponding to 5.7 events per parameter. The overall spline test in EOCRC was statistically significant (p = 0.048), while the categorical and linear models were not. Given the limited number of events, the greater flexibility of the spline model may have contributed to this finding through overfitting.

Another limitation was missing BMI data, primarily among patients who underwent emergency surgery, for whom height and weight were often not recorded. Many of these patients had stage IV disease or underwent non-radical surgery and would therefore have been excluded for other reasons. Among radically operated patients, recurrence rates were similar regardless of BMI data availability. Nevertheless, selection bias due to missing BMI data cannot be excluded.

Categorising BMI also has limitations. The underweight and obese EOCRC groups were small, with only 3 and 15 patients, respectively. Most patients in the obesity category had a BMI just above 30, and patients with BMI above 40 were excluded. The findings may therefore not apply to patients with more severe obesity.

Collider bias and residual confounding may also have influenced the results. Since the study included patients with CRC, selection on having developed the disease may have introduced associations between BMI and other risk factors, such as smoking [31]. Smoking is associated with lower BMI and worse prognosis. This could contribute to an apparent protective association of higher BMI. Smoking, diet, comorbidities and molecular tumour characteristics were not available for this analysis.

## Conclusions

BMI at diagnosis was associated with recurrence in EOCRC, with the lowest hazards in the overweight range, whereas no association was observed in AOCRC. The direction of the association was consistent across sensitivity analyses, including the exclusion of patients with stage IV disease. The observed pattern may be consistent with an obesity paradox in EOCRC. However, evidence for nonlinearity was inconclusive, the difference between the cohorts was not statistically significant, and prediagnostic weight loss could not be accounted for. Larger studies incorporating prediagnostic weight and body composition are needed to confirm these findings and explore potential explanations.

## Data Availability

The research database was approved by the Regional Ethics Review Board (no. 118-15), and all patients provided informed consent for inclusion.

## Abbreviations

BMI: Body Mass Index
CRC: Colorectal Cancer
EOCRC: Early-Onset Colorectal Cancer
AOCRC: Average-Onset Colorectal Cancer
HR: Hazard Ratio
LRT: Likelihood-ratio Test
CI: Confidence Interval
SD: Standard Deviation
AJCC: American Joint Committee on Cancer
EMVI: Extramural Vascular Invasion

## Data Availability

The datasets analysed during the current study are not available for sharing due to patient confidentiality and restrictions under the current ethical approval.

