## Supplemental File for "Weighing the Hazards: Body Mass Index and Recurrence in Early-Onset Colorectal Cancer"

**Supplement tables**

**eTable 1.** Demographic, clinical and histopathological characteristics by BMI category in the EOCRC cohort

| | *Underweight (n=3)* | *Normal weight (n=79)* | *Overweight (n=61)* | *Obese (n=15)* | *Total (n=158)* | *P-value* |
|---|---|---|---|---|---|---|
| | | | ***Demographic*** | | | |
| **Age, years** | 34.3 ± 8.3 (25-41) | 42.0 ± 6.1 (24-49) | 44.4 ± 4.7 (27-49) | 44.3 ± 4.3 (35-49) | 43.0 ± 5.7 (24-49) | **0.003** |
| **Sex** | | | | | | 0.08 |
| Male | 0 (0.0) | 37 (46.8) | 36 (59.0) | 10 (66.7) | 83 (52.5) | |
| Female | 3 (100.0) | 42 (53.2) | 25 (41.0) | 5 (33.3) | 75 (47.5) | |
| | | | ***Disease characteristics*** | | | |
| **Tumour location** | | | | | | 0.24 |
| Right colon | 1 (33.3) | 25 (31.6) | 19 (31.1) | 7 (46.7) | 52 (32.9) | |
| Left colon | 0 (0.0) | 24 (30.4) | 10 (16.4) | 4 (26.7) | 38 (24.1) | |
| Rectum | 2 (66.7) | 30 (38.0) | 32 (52.5) | 4 (26.7) | 68 (43.0) | |
| **Stage** | | | | | | **0.02** |
| I | 0 (0.0) | 12 (15.2) | 11 (18.0) | 2 (13.3) | 25 (15.8) | |
| II | 1 (33.3) | 18 (22.8) | 17 (27.9) | 0 (0.0) | 36 (22.8) | |
| III | 1 (33.3) | 34 (43.0) | 32 (52.5) | 11 (73.3) | 78 (49.4) | |
| IV | 1 (33.3) | 13 (16.5) | 1 (1.6) | 2 (13.3) | 17 (10.8) | |
| ypT0N0 | 0 (0.0) | 2 (2.5) | 0 (0.0) | 0 (0.0) | 2 (1.3) | |
| **T stage** | | | | | | 0.62 |
| T0 | 0 (0.0) | 2 (2.5) | 0 (0.0) | 1 (6.7) | 3 (1.9) | |
| T1 | 0 (0.0) | 4 (5.1) | 2 (3.3) | 1 (6.7) | 7 (4.4) | |
| T2 | 0 (0.0) | 12 (15.2) | 16 (26.2) | 3 (20.0) | 31 (19.6) | |
| T3 | 3 (100.0) | 45 (57.0) | 36 (59.0) | 9 (60.0) | 93 (58.9) | |
| T4 | 0 (0.0) | 15 (19.0) | 7 (11.5) | 1 (6.7) | 23 (14.6) | |
| TX | 0 (0.0) | 1 (1.3) | 0 (0.0) | 0 (0.0) | 1 (0.6) | |
| **N stage** | | | | | | **0.008** |
| N0 | 2 (66.7) | 34 (43.0) | 30 (49.2) | 3 (20.0) | 69 (43.7) | |
| N1 | 0 (0.0) | 17 (21.5) | 24 (39.3) | 8 (53.3) | 49 (31.0) | |
| N2 | 1 (33.3) | 25 (31.6) | 7 (11.5) | 4 (26.7) | 37 (23.4) | |
| NX | 0 (0.0) | 3 (3.8) | 0 (0.0) | 0 (0.0) | 3 (1.9) | |
| **Differentiation** | | | | | | 0.40 |
| Poor | 1 (33.3) | 23 (29.1) | 14 (23.0) | 6 (40.0) | 44 (27.8) | |
| Moderately | 2 (66.7) | 38 (48.1) | 39 (63.9) | 8 (53.3) | 87 (55.1) | |
| Well | 0 (0.0) | 3 (3.8) | 1 (1.6) | 1 (6.7) | 5 (3.2) | |
| Mucinous | 0 (0.0) | 10 (12.7) | 7 (11.5) | 0 (0.0) | 17 (10.8) | |
| Not reported | 0 (0.0) | 5 (6.3) | 0 (0.0) | 0 (0.0) | 5 (3.2) | |
| **Tumour perforation (n=56)** | | | | | | 0.62 |

| | *Underweight (n=3)* | *Normal weight (n=79)* | *Overweight (n=61)* | *Obese (n=15)* | *Total (n=158)* | *P-value* |
|---|---|---|---|---|---|---|
| No | 1 (100.0) | 21 (87.5) | 24 (96.0) | 6 (100.0) | 52 (92.9) | |
| Yes | 0 (0.0) | 3 (12.5) | 1 (4.0) | 0 (0.0) | 4 (7.1) | |
| Missing | | | | | 102 | |
| **Perineural growth (n=57)** | | | | | | 0.38 |
| No | 1 (100.0) | 21 (84.0) | 23 (92.0) | 4 (66.7) | 49 (86.0) | |
| Yes | 0 (0.0) | 4 (16.0) | 2 (8.0) | 2 (33.3) | 8 (14.0) | |
| Missing | | | | | 101 | |
| **Vascular growth (n=57)** | | | | | | 0.35 |
| No | 1 (100.0) | 13 (52.0) | 18 (72.0) | 3 (50.0) | 35 (61.4) | |
| Yes | 0 (0.0) | 12 (48.0) | 7 (28.0) | 3 (50.0) | 22 (38.6) | |
| Missing | | | | | 101 | |
| **EMVI (n=39)** | | | | | | 0.30 |
| No | 1 (100.0) | 9 (52.9) | 13 (81.2) | 3 (60.0) | 26 (66.7) | |
| Yes | 0 (0.0) | 8 (47.1) | 3 (18.8) | 2 (40.0) | 13 (33.3) | |
| Missing | | | | | 119 | |
| ***Treatment characteristics*** | | | | | | |
| **Neoadjuvant radiotherapy, rectal cancer only (n=68)** | | | | | | 0.50 |
| No | 0 (0.0) | 11 (36.7) | 11 (34.4) | 0 (0.0) | 22 (32.4) | |
| Yes | 2 (100.0) | 19 (63.3) | 21 (65.6) | 4 (100.0) | 46 (67.6) | |
| **Neoadjuvant chemotherapy** | | | | | | 0.21 |
| No | 2 (66.7) | 68 (86.1) | 57 (93.4) | 14 (93.3) | 141 (89.2) | |
| Yes | 1 (33.3) | 11 (13.9) | 4 (6.6) | 1 (6.7) | 17 (10.8) | |
| **Operation type** | | | | | | 0.053 |
| Right colon resection | 1 (33.3) | 15 (19.0) | 16 (26.2) | 5 (33.3) | 37 (23.4) | |
| Left colon resection | 0 (0.0) | 17 (21.5) | 10 (16.4) | 4 (26.7) | 31 (19.6) | |
| Rectal resection | 2 (66.7) | 29 (36.7) | 33 (54.1) | 4 (26.7) | 68 (43.0) | |
| (Procto)colectomy | 0 (0.0) | 9 (11.4) | 2 (3.3) | 1 (6.7) | 12 (7.6) | |
| Other | 0 (0.0) | 9 (11.4) | 0 (0.0) | 1 (6.7) | 10 (6.3) | |
| **Elective surgery** | | | | | | 0.19 |
| No | 1 (33.3) | 3 (3.8) | 2 (3.3) | 0 (0.0) | 6 (3.8) | |
| Yes | 2 (66.7) | 76 (96.2) | 59 (96.7) | 15 (100.0) | 152 (96.2) | |
| **Laparoscopic surgery (n=156)** | | | | | | 0.11 |
| No | 3 (100.0) | 57 (74.0) | 36 (59.0) | 8 (53.3) | 104 (66.7) | |
| Yes | 0 (0.0) | 20 (26.0) | 25 (41.0) | 7 (46.7) | 52 (33.3) | |
| Missing | | | | | 2 | |
| **Radical resection** | | | | | | 0.12 |
| At primary operation | 2 (66.7) | 71 (89.9) | 59 (96.7) | 14 (93.3) | 146 (92.4) | |

| | *Underweight (n=3)* | *Normal weight (n=79)* | *Overweight (n=61)* | *Obese (n=15)* | *Total (n=158)* | *P-value* |
|---|---|---|---|---|---|---|
| After additional surgery | 1 (33.3) | 8 (10.1) | 2 (3.3) | 1 (6.7) | 12 (7.6) | |
| **Resection of metastases (n=83)** | | | | | | **0.03** |
| No | 2 (100.0) | 29 (80.6) | 35 (100.0) | 9 (90.0) | 75 (90.4) | |
| Yes | 0 (0.0) | 7 (19.4) | 0 (0.0) | 1 (10.0) | 8 (9.6) | |
| Missing | | | | | 75 | |
| **Adjuvant chemotherapy** | | | | | | 0.08 |
| No | 0 (0.0) | 21 (26.6) | 25 (41.0) | 2 (13.3) | 48 (30.4) | |
| Yes | 3 (100.0) | 58 (73.4) | 36 (59.0) | 13 (86.7) | 110 (69.6) | |
| **Complementary operation** | | | | | | 0.12 |
| No | 2 (66.7) | 71 (89.9) | 59 (96.7) | 14 (93.3) | 146 (92.4) | |
| Yes | 1 (33.3) | 8 (10.1) | 2 (3.3) | 1 (6.7) | 12 (7.6) | |
| | | | ***Outcomes*** | | | |
| **New primary colorectal cancer** | | | | | | 0.11 |
| No | 3 (100.0) | 79 (100.0) | 61 (100.0) | 14 (93.3) | 157 (99.4) | |
| Yes | 0 (0.0) | 0 (0.0) | 0 (0.0) | 1 (6.7) | 1 (0.6) | |
| **Recurrence** | | | | | | 0.11 |
| No | 2 (66.7) | 53 (67.1) | 51 (83.6) | 12 (80.0) | 118 (74.7) | |
| Yes | 1 (33.3) | 26 (32.9) | 10 (16.4) | 3 (20.0) | 40 (25.3) | |

Values are n (%) unless stated otherwise; percentages are of patients with data on the variable. Age is mean ± SD (range). BMI categories: underweight < 18.5, normal weight 18.5-24.9, overweight 25-29.9 and obese 30-40; patients with BMI above 40 were excluded. P-values: ANOVA for age and Fisher's exact test for categorical variables, excluding missing values.

ypT0N0, complete pathological response after neoadjuvant treatment; EMVI, extramural vascular invasion. Neoadjuvant radiotherapy is reported for patients with rectal cancer only.

**eTable 2.** Demographic, clinical and histopathological characteristics by BMI category in the AOCRC cohort

| | *Underweight (n=32)* | *Normal weight (n=744)* | *Overweight (n=627)* | *Obese (n=260)* | *Total (n=1663)* | *P-value* |
|---|---|---|---|---|---|---|
| | | | ***Demographic*** | | | |
| **Age, years** | 71.6 ± 10.8 (50-91) | 72.5 ± 9.9 (50-93) | 70.3 ± 9.3 (50-95) | 69.0 ± 9.1 (50-90) | 71.1 ± 9.7 (50-95) | **<0.001** |
| **Sex** | | | | | | **<0.001** |
| Male | 2 (6.2) | 365 (49.1) | 385 (61.4) | 137 (52.7) | 889 (53.5) | |
| Female | 30 (93.8) | 379 (50.9) | 242 (38.6) | 123 (47.3) | 774 (46.5) | |
| | | | ***Disease characteristics*** | | | |
| **Tumour location** | | | | | | **0.046** |
| Right colon | 13 (40.6) | 275 (37.0) | 186 (29.7) | 76 (29.2) | 550 (33.1) | |
| Left colon | 7 (21.9) | 194 (26.1) | 177 (28.2) | 84 (32.3) | 462 (27.8) | |
| Rectum | 12 (37.5) | 275 (37.0) | 264 (42.1) | 100 (38.5) | 651 (39.1) | |
| **Stage** | | | | | | 0.80 |
| I | 5 (15.6) | 171 (23.0) | 139 (22.2) | 63 (24.2) | 378 (22.7) | |
| II | 15 (46.9) | 243 (32.7) | 188 (30.0) | 86 (33.1) | 532 (32.0) | |
| III | 10 (31.2) | 270 (36.3) | 251 (40.0) | 91 (35.0) | 622 (37.4) | |
| IV | 2 (6.2) | 55 (7.4) | 42 (6.7) | 18 (6.9) | 117 (7.0) | |
| ypT0N0 | 0 (0.0) | 5 (0.7) | 7 (1.1) | 2 (0.8) | 14 (0.8) | |
| **T stage** | | | | | | 0.20 |
| T0 | 0 (0.0) | 8 (1.1) | 9 (1.4) | 3 (1.2) | 20 (1.2) | |
| T1 | 2 (6.2) | 41 (5.5) | 55 (8.8) | 24 (9.2) | 122 (7.3) | |
| T2 | 8 (25.0) | 175 (23.5) | 130 (20.7) | 57 (21.9) | 370 (22.2) | |
| T3 | 13 (40.6) | 395 (53.1) | 323 (51.5) | 141 (54.2) | 872 (52.4) | |
| T4 | 8 (25.0) | 123 (16.5) | 107 (17.1) | 33 (12.7) | 271 (16.3) | |
| TX | 1 (3.1) | 2 (0.3) | 3 (0.5) | 2 (0.8) | 8 (0.5) | |
| **N stage** | | | | | | 0.37 |
| N0 | 20 (62.5) | 421 (56.6) | 331 (52.8) | 149 (57.3) | 921 (55.4) | |
| N1 | 11 (34.4) | 226 (30.4) | 189 (30.1) | 71 (27.3) | 497 (29.9) | |
| N2 | 1 (3.1) | 90 (12.1) | 96 (15.3) | 36 (13.8) | 223 (13.4) | |
| NX | 0 (0.0) | 7 (0.9) | 11 (1.8) | 4 (1.5) | 22 (1.3) | |
| **Differentiation** | | | | | | 0.87 |
| Poor | 10 (31.2) | 169 (22.7) | 136 (21.7) | 50 (19.2) | 365 (21.9) | |
| Moderately | 17 (53.1) | 467 (62.8) | 390 (62.2) | 161 (61.9) | 1035 (62.2) | |
| Well | 2 (6.2) | 26 (3.5) | 26 (4.1) | 14 (5.4) | 68 (4.1) | |
| Mucinous | 3 (9.4) | 70 (9.4) | 62 (9.9) | 30 (11.5) | 165 (9.9) | |
| Not reported | 0 (0.0) | 12 (1.6) | 13 (2.1) | 5 (1.9) | 30 (1.8) | |

| | *Underweight (n=32)* | *Normal weight (n=744)* | *Overweight (n=627)* | *Obese (n=260)* | *Total (n=1663)* | *P-value* |
|---|---|---|---|---|---|---|
| **Tumour perforation (n=682)** | | | | | | 0.59 |
| No | 12 (92.3) | 273 (95.1) | 255 (96.2) | 109 (93.2) | 649 (95.2) | |
| Yes | 1 (7.7) | 14 (4.9) | 10 (3.8) | 8 (6.8) | 33 (4.8) | |
| Missing | | | | | 981 | |
| **Perineural growth (n=662)** | | | | | | 0.60 |
| No | 10 (76.9) | 244 (87.5) | 222 (86.4) | 101 (89.4) | 577 (87.2) | |
| Yes | 3 (23.1) | 35 (12.5) | 35 (13.6) | 12 (10.6) | 85 (12.8) | |
| Missing | | | | | 1001 | |
| **Vascular growth (n=680)** | | | | | | 0.75 |
| No | 9 (69.2) | 191 (66.6) | 186 (69.9) | 74 (64.9) | 460 (67.6) | |
| Yes | 4 (30.8) | 96 (33.4) | 80 (30.1) | 40 (35.1) | 220 (32.4) | |
| Missing | | | | | 983 | |
| **EMVI (n=468)** | | | | | | 0.54 |
| No | 5 (62.5) | 140 (69.0) | 133 (75.1) | 58 (72.5) | 336 (71.8) | |
| Yes | 3 (37.5) | 63 (31.0) | 44 (24.9) | 22 (27.5) | 132 (28.2) | |
| Missing | | | | | 1195 | |
| | | | ***Treatment characteristics*** | | | |
| **Neoadjuvant radiotherapy, rectal cancer only (n=650)** | | | | | | 0.73 |
| No | 4 (33.3) | 114 (41.5) | 117 (44.5) | 46 (46.0) | 281 (43.2) | |
| Yes | 8 (66.7) | 161 (58.5) | 146 (55.5) | 54 (54.0) | 369 (56.8) | |
| Missing | | | 1 | | | |
| **Neoadjuvant chemotherapy** | | | | | | 0.35 |
| No | 29 (90.6) | 708 (95.2) | 591 (94.3) | 251 (96.5) | 1579 (94.9) | |
| Yes | 3 (9.4) | 36 (4.8) | 36 (5.7) | 9 (3.5) | 84 (5.1) | |
| **Operation type** | | | | | | **0.02** |
| Right colon resection | 12 (37.5) | 243 (32.7) | 170 (27.1) | 68 (26.2) | 493 (29.6) | |
| Left colon resection | 3 (9.4) | 170 (22.8) | 147 (23.4) | 72 (27.7) | 392 (23.6) | |
| Rectal resection | 14 (43.8) | 287 (38.6) | 283 (45.1) | 106 (40.8) | 690 (41.5) | |
| (Procto)colectomy | 1 (3.1) | 13 (1.7) | 11 (1.8) | 10 (3.8) | 35 (2.1) | |
| Other | 2 (6.2) | 31 (4.2) | 16 (2.6) | 4 (1.5) | 53 (3.2) | |
| **Elective surgery** | | | | | | 0.96 |
| No | 1 (3.1) | 31 (4.2) | 28 (4.5) | 10 (3.8) | 70 (4.2) | |
| Yes | 31 (96.9) | 713 (95.8) | 599 (95.5) | 250 (96.2) | 1593 (95.8) | |
| **Laparoscopic surgery (n=1641)** | | | | | | **0.005** |

| | *Underweight (n=32)* | *Normal weight (n=744)* | *Overweight (n=627)* | *Obese (n=260)* | *Total (n=1663)* | *P-value* |
|---|---|---|---|---|---|---|
| No | 28 (87.5) | 476 (64.5) | 371 (60.5) | 176 (68.2) | 1051 (64.0) | |
| Yes | 4 (12.5) | 262 (35.5) | 242 (39.5) | 82 (31.8) | 590 (36.0) | |
| Missing | | | | | 22 | |
| **Radical resection** | | | | | | 0.41 |
| At primary operation | 29 (90.6) | 713 (95.8) | 593 (94.6) | 249 (95.8) | 1584 (95.2) | |
| After additional surgery | 3 (9.4) | 31 (4.2) | 34 (5.4) | 11 (4.2) | 79 (4.8) | |
| **Resection of metastases (n=901)** | | | | | | 0.37 |
| No | 19 (100.0) | 349 (92.8) | 341 (95.3) | 139 (93.9) | 848 (94.1) | |
| Yes | 0 (0.0) | 27 (7.2) | 17 (4.7) | 9 (6.1) | 53 (5.9) | |
| Missing | | | | | 762 | |
| **Adjuvant chemotherapy** | | | | | | **0.002** |
| No | 24 (75.0) | 502 (67.5) | 363 (57.9) | 163 (62.7) | 1052 (63.3) | |
| Yes | 8 (25.0) | 242 (32.5) | 264 (42.1) | 97 (37.3) | 611 (36.7) | |
| **Complementary operation (n=1662)** | | | | | | 0.40 |
| No | 29 (90.6) | 711 (95.6) | 588 (93.9) | 247 (95.0) | 1575 (94.8) | |
| Yes | 3 (9.4) | 33 (4.4) | 38 (6.1) | 13 (5.0) | 87 (5.2) | |
| Missing | | | | | 1 | |
| ***Outcomes*** | | | | | | |
| **New primary colorectal cancer** | | | | | | 0.51 |
| No | 31 (96.9) | 738 (99.2) | 620 (98.9) | 256 (98.5) | 1645 (98.9) | |
| Yes | 1 (3.1) | 6 (0.8) | 7 (1.1) | 4 (1.5) | 18 (1.1) | |
| **Recurrence** | | | | | | 0.93 |
| No | 26 (81.2) | 621 (83.5) | 518 (82.6) | 213 (81.9) | 1378 (82.9) | |
| Yes | 6 (18.8) | 123 (16.5) | 109 (17.4) | 47 (18.1) | 285 (17.1) | |

Values are n (%) unless stated otherwise; percentages are of patients with data on the variable. Age is mean ± SD (range). BMI categories: underweight < 18.5, normal weight 18.5-24.9, overweight 25-29.9 and obese 30-40; patients with BMI above 40 were excluded. P-values: ANOVA for age and the chi-squared test for categorical variables, excluding missing values.

ypT0N0, complete pathological response after neoadjuvant treatment; EMVI, extramural vascular invasion. Neoadjuvant radiotherapy is reported for patients with rectal cancer only.

**eTable 3.** Site of recurrence by BMI category in EOCRC and AOCRC

| | *Underweight* | *Normal weight* | *Overweight* | *Obese* | *Total* |
|---|---|---|---|---|---|
| | | | **EOCRC** | | |
| Patients, n | 3 | 79 | 61 | 15 | 158 |
| **Recurrence** | 1 (33.3) | 26 (32.9) | 10 (16.4) | 3 (20.0) | 40 (25.3) |
| **Locoregional** | 0 (0.0) | 6 (7.6) | 0 (0.0) | 2 (13.3) | 8 (5.1) |
| Local | 0 (0.0) | 4 (5.1) | 0 (0.0) | 1 (6.7) | 5 (3.2) |
| Regional lymph nodes | 0 (0.0) | 2 (2.5) | 0 (0.0) | 1 (6.7) | 3 (1.9) |
| **Distant** | 1 (33.3) | 20 (25.3) | 10 (16.4) | 1 (6.7) | 32 (20.3) |
| Liver | 0 (0.0) | 7 (8.9) | 3 (4.9) | 1 (6.7) | 11 (7.0) |
| Lung | 1 (33.3) | 3 (3.8) | 3 (4.9) | 0 (0.0) | 7 (4.4) |
| Peritoneum | 0 (0.0) | 3 (3.8) | 1 (1.6) | 0 (0.0) | 4 (2.5) |
| Multiple sites | 0 (0.0) | 5 (6.3) | 3 (4.9) | 0 (0.0) | 8 (5.1) |
| Other | 0 (0.0) | 2 (2.5) | 0 (0.0) | 0 (0.0) | 2 (1.3) |
| | | | **AOCRC** | | |
| Patients, n | 32 | 744 | 627 | 260 | 1663 |
| **Recurrence** | 6 (18.8) | 123 (16.5) | 109 (17.4) | 47 (18.1) | 285 (17.1) |
| **Locoregional** | 2 (6.2) | 18 (2.4) | 9 (1.4) | 6 (2.3) | 35 (2.1) |
| Local | 1 (3.1) | 15 (2.0) | 2 (0.3) | 2 (0.8) | 20 (1.2) |
| Regional lymph nodes | 1 (3.1) | 3 (0.4) | 7 (1.1) | 4 (1.5) | 15 (0.9) |
| **Distant** | 4 (12.5) | 105 (14.1) | 100 (15.9) | 41 (15.8) | 250 (15.0) |
| Liver | 0 (0.0) | 31 (4.2) | 28 (4.5) | 17 (6.5) | 76 (4.6) |
| Lung | 1 (3.1) | 26 (3.5) | 25 (4.0) | 9 (3.5) | 61 (3.7) |
| Peritoneum | 1 (3.1) | 4 (0.5) | 6 (1.0) | 1 (0.4) | 12 (0.7) |
| Multiple sites | 2 (6.2) | 37 (5.0) | 38 (6.1) | 12 (4.6) | 89 (5.4) |
| Other | 0 (0.0) | 7 (0.9) | 3 (0.5) | 2 (0.8) | 12 (0.7) |

Values are n (%) unless stated otherwise. Multiple sites are multiple distant metastatic sites; other sites are bone, brain, ovary, subcutaneous tissue and other locations.

**eFig 1** Kaplan Meier curve in early-onset colorectal cancer

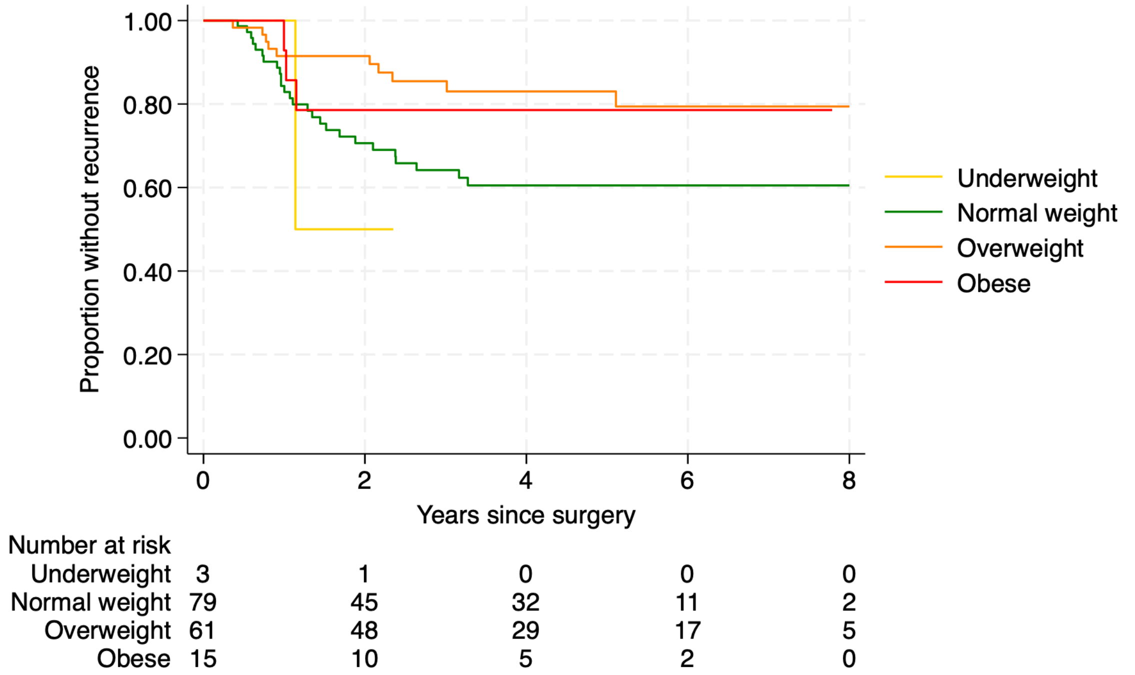


**eFig 1** Kaplan-Meier curve showing the proportion of patients without recurrence by BMI category in early-onset colorectal cancer (EOCRC, n = 158). Time was measured from surgery.

**eFig 2** Kaplan Meier estimate in average-onset colorectal cancer

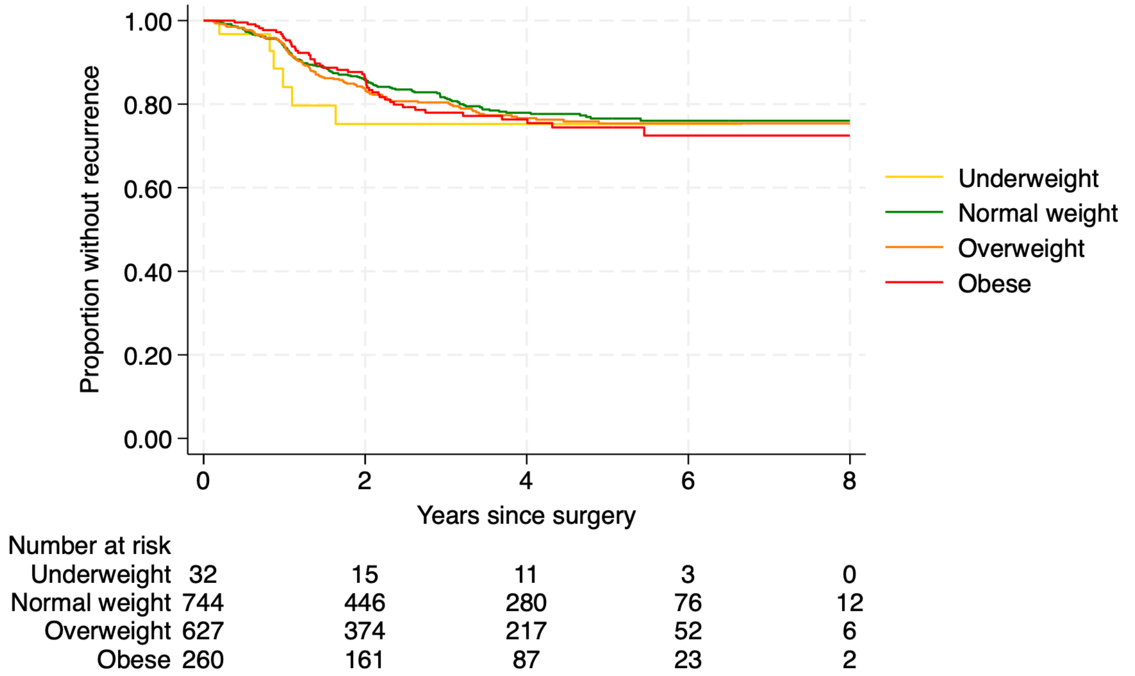


**eFig 2** Kaplan-Meier curve showing the proportion of patients without recurrence by BMI category in average-onset colorectal cancer (AOCRC, n = 1,663). Time was measured from surgery.
